# Deep learning-based computer-aided diagnostic models *versus* other methods for predicting malignancy risk in CT-detected pulmonary nodules

**DOI:** 10.1101/2023.06.06.23291012

**Authors:** Wahyu Wulaningsih, Abdullah Akram, Janella Benemile, Ruth Kathyrn, Filippo Croce, Johnathan Watkins

## Abstract

**Importance:** There has been growing interest in the use of artificial intelligence (deep learning) to help achieve early diagnosis of prevalent diseases. None moreso than in lung cancer, where a combination of factors, including the high prevalence of nodules, the low prevalence of malignant nodules, and the indeterminacy of many nodules mean that it is fertile ground for the deployment of accurate, high-throughput deep learning (DL)-based tools.

**Objective:** To survey the landscape of externally validated DL-based computer-aided diagnostic (CADx) models, and assess their diagnostic performance for predicting the risk of malignancy in computed tomography (CT)-detected pulmonary nodules.

**Data sources:** An electronic search was performed in the MEDLINE (PubMed), EMBASE, Science Citation Index, Cochrane Library databases (from inception to 10 April 2023).

**Study selection:** Studies were deemed eligible if they were peer-reviewed experimental or observational articles that analysed the diagnostic performance of externally validated DL-based CADx models for the prediction of malignancy risk, with a direct comparison to models widely used in clinical practice.

**Data extraction and synthesis:** PRISMA guidelines were followed for the identification, screening, and selection process. A bivariate random-effect approach for the meta-analysis on the included studies was used. Quality Assessment of Diagnosis Accuracy Studies 2 (QUADAS-2) was used to assess risk of bias and applicability.

**Main outcomes and measures:** Main outcomes included sensitivity, specificity, and area under the curve (AUC).

**Results:** After screening, 20 studies were included, comprising 7,664 participants and 10,128 nodules, of which 2,126 were malignant. DL-based CADx models were 15.8% more sensitive than physician judgement alone, and 35.4% more than clinical risk models alone. They had a similar pooled specificity as physician judgement alone (0.77 [95% CI: 0.69 –0.84] *v* 0.80 [95% CI: 0.71 –0.86], respectively), but were 5.5% more specific than clinical risk models alone. Accounting for threshold effects, DL-based CADx models had superior summary areas under the receiver operating characteristic curve (sAUROC), with relative sAUROCs of 1.06 (95% CI: 1.03–1.08) and 1.22 (95% CI: 1.19–1.24), as compared to physician judgement and clinical risk models alone, respectively.

**Conclusions and relevance:** DL-based models show superior or comparable diagnostic performance when externally validated against widely used methods, such as the Brock and Mayo models. They have the potential to fulfil an unmet clinical-management need alongside experienced physician image readers. The included studies reported a high degree of heterogeneity, with threshold effects particularly prominent. Future research may consider more prospective studies and human-experimental studies.

**Key points:** *Question:* How effective are image-based, computer-aided diagnostic models that use deep learning methods to predict the malignancy risk of pulmonary nodules as compared with other methods used in clinical practice?

*Findings:* This systematic review and meta-analysis identified 20 observational studies (7,664 participants; 10,128 pulmonary nodules) from which pooled analyses found deep learning-based models to have a sensitivity of 0.88, specificity of 0.77, and summary area under the curve of 0.90 in predicting malignancy in pulmonary nodules. This was superior or comparable to other methods routinely used in clinical practice.

*Meaning:* Deep learning-based models are already being used in clinical practice in certain settings for nodule management. The results show their diagnostic performance justifies wider and more routine deployment.

## Introduction

Five-year survival rates in the US for lung cancer fall from 73% at stage I to just 9–13% at stages IIIB and IV.^1^ Hence, diagnosing lung cancer early is critical to reducing lung cancer mortality rates. Lung cancer is predominantly asymptomatic in its early stages, with pulmonary nodules often being the first sign.^2^ Pulmonary nodules are discrete lung lesions, measuring ≤30 mm in size (average of axial diameters). Approximately 5% of these nodules are malignant.^3^ Nodules, both benign and malignant, are detected in approximately 1.6 million people in the US each year.^3^ The majority of these are detected by computed tomography (CT) scans, of which more than 12 million are performed in the US each year.^4^ These facts combine to show that the detection and discrimination of pulmonary nodules are the most important means of diagnosing lung cancer early, and CT scans the most important modality.

Pulmonary nodules are easy to detect, but difficult to discriminate. Assessing the risk of indeterminate nodules – nodules without obvious signs of benignity (such as calcification) or obvious signs of malignancy (such as spiculation) – pose a particular challenge.^5,6^ Multiple clinical risk prediction models are used to aid the physician in assesing nodules in order to diagnose lung cancer or refer high-risk cases for further, more invasive investigation. Most validated clinical risk models apply multivariable logistic regression to clinico-demographic predictors (such as age, family history, and smoking history) and radiological predictors (such as nodule size, morphology, and location). Externally validated models in clinical use include the Mayo model, Brock (or PanCan) model, and the Peking University People’s Hospital (PKUPH) model.^7–9^

Recently, image-based artificial intelligence (AI) models that use deep learning (DL) methods have emerged to predict this malignancy risk.^10^ One of the advantages of image-based computer-aided diagnostic (CADx) models is their ease and speed of use *versus* traditional risk tools, which require manual entry of input data. This manual entry leads to low adoption rates, which may fall even further as the number of patients entering the care pathway increases. As such, adding DL capability to image-based CADx models has the potential to fulfil an unmet clinical-management need, providing they produce comparable diagnostic performance.

The objective of this systematic review and meta-analysis was to survey the landscape of externally validated DL-based CADx models, and assess their diagnostic performance for predicting the risk of malignancy in CT-detected pulmonary nodules. Two previous systematic reviews have been conducted on studies of DL-based CADx models that diagnose lung cancer from pulmonary nodules,^11,12^ but none have conducted pooled analysis on models that have been externally validated against models currently used in clinical practice. Some degree of external validation in populations other than the population used to train the new model is essential to ensuring they are sufficiently robust to training bias. This is the first systematic review to provide such a pooled analysis of studies, in that it considers only those studies that directly compare DL-based CADx models with physician judgement, clinical risk models, or Lung-RADS-based models.

## Methods

### Search strategy and screening

An electronic search was performed in the MEDLINE (PubMed), EMBASE, Science Citation Index, and Cochrane Library databases (from inception to 10 April 2023). Relevant English-language studies only were sought. Duplicate studies, case reports & series, non-systematic review articles, non-peer reviewed studies, non-human studies, meeting abstracts & proceedings, and unpublished studies were all excluded. The full set of keyword search terms may be found in eTable 1 in the Supplementary Material. Reference lists of key studies and domain-related systematic reviews were investigated for further studies that the search may have missed. This study followed the Preferred Reporting Items for Systematic Reviews and Meta-analyses (PRISMA) reporting guidelines.^13^

A total of 7,116 studies were found after removing duplicates (Figure 1). After screening out ineligible studies from their title and abstract, the full text of 69 studies were retrieved for final screening. Two reviewers (JB and RK) independently reviewed each text.

**Figure 1.**
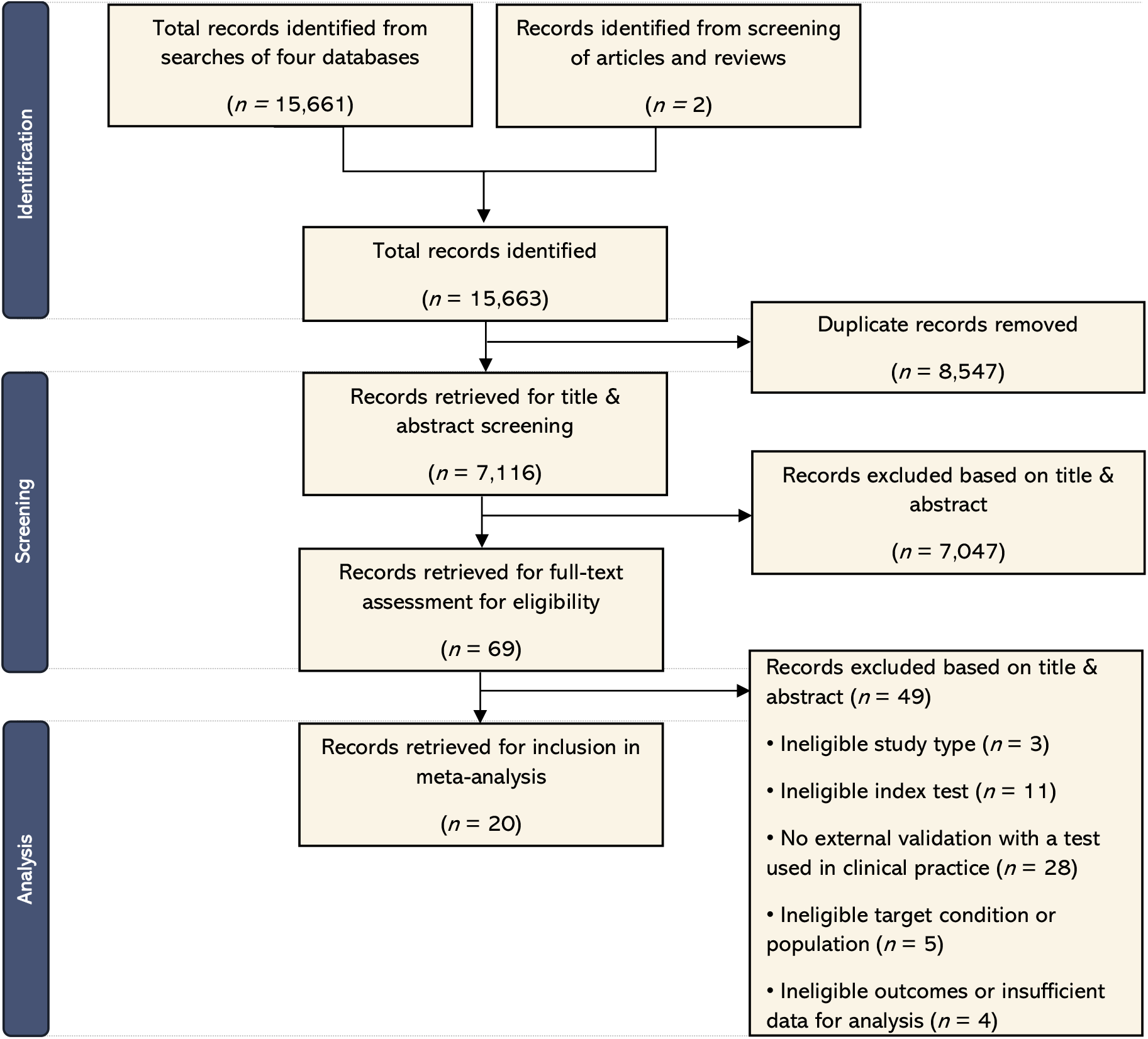
Literature search flow diagram

### Data extraction and quality assessment

From the included studies, the following information was extracted and tabulated independently by two reviewers (JB and RK): author; publication date; funding source; study type; study population country(ies); setting; outcome type(s); index test(s); reference test; number of participants in each validation dataset; number of nodules in each validation dataset; prevalence of malignancy among nodules; age range; sex; patient exclusions; proportion of smokers (current and former); nodule diameter range; median nodule diameter; nodule type(s); threshold (operating cut-off point); route of detection; and the outcomes reported. The data were subsequently checked for quality (AA, WW, and JW). Studies that met the following criteria were included:

- ***Study type***. Studies should be human experimental studies or human observational studies
- ***Index tests***.
  - Index test being described and investigated should use AI or DL methods (DL-based model) – defined as the self-reported use of AI or DL – to classify or otherwise predict the risk of malignancy in pulmonary nodules detected via CT scans
  - External validation of the DL-based CADx model should be performed on data not used for the initial development of the DL-based CADx model and compared with other methods that are in widespread clinical use, the categories of which are:
    - Physician judgement (radiological image readers)
    - Clinical risk models (multivariable statistical models that use clinico-demographic and radiological variables as inputs)
    - Lung-RADS-based models (that allow computers or humans to automatically classify nodules based on nodule size, type, and stability over time)^14^
- ***Reference tests***. Studies should confirm malignancy diagnosis via histopathological (biopsy) within the follow-up period after initial nodule detection
- ***Target condition and population***. Study participants should be ≥18 years old, with at least one solid or part-solid pulmonary nodule (0–30 mm), as identified via CT scan (i.e. studies on ground-glass nodules [GGNs] only are excluded)
- ***Outcomes***. Studies should report at least one of: sensitivity, specificity; areas under the curve (AUC); diagnostic odds ratios; or the number of true-positive, false-negative, true-negative, or false-positive cases (as confirmed by histopathological analysis)

Risk of bias and applicability was independently assessed by AA, JB, and JW using the Quality Assessment of Diagnostic Studies 2 (QUADAS-2) tool (eFigure 1 in the Supplementary Material).^15^

### Statistical analysis and quantitative synthesis

A meta-analysis of all included studies reporting diagnostic performance outcomes was conducted. For each of the index test types (DL-based CADx models; physician judgement alone; clinical risk models alone; Lung-RADS-based models alone), pooled estimates of sensitivity, specificity, and AUC were calculated using a bivariate, random-effects approach, along with their respective 95% confidence intervals (CIs). Summary AUROC curves were plotted.

To assess heterogeneity and inconsistency among the studies, χ^2^ statistic and I^2^ index values were calculated. An I^2^ value greater than 75% was considered indicative of substantial heterogeneity.

The Deeks funnel plot asymmetry test was performed to test for publication bias. A two-sided p < 0.05 result was assumed to be statistically significant.

For the data available for extraction, we explored the sensitivity of the main pooled estimates to the study and population-outcome characteristics by conducting sub-group analyses. Once again, pooled sensitivity, specificity, and AUC estimates, along with χ^2^ statistic and I^2^ index values, were calculated. The data were stratified by sub-group such as prevalence, route of detection, and median nodule size. This also helped uncover sources of heterogeneity.

Review Manager (RevMan) version 5.4 and Stata software, version 18 (StataCorp LLC) were used to conduct the statistical analyses.^16,17^

## Results

### Study characteristics

The literature search identified 20 studies for inclusion (Figure 1), comprising 38 validation datasets, representing 7,664 participants and 10,128 pulmonary nodules. Of these nodules, 2,126 were confirmed to be malignant (histopathological ground truth) within the follow-up period (on average, 24 months). A summary of the included studies is provided in Table 1.

**Table 1.**
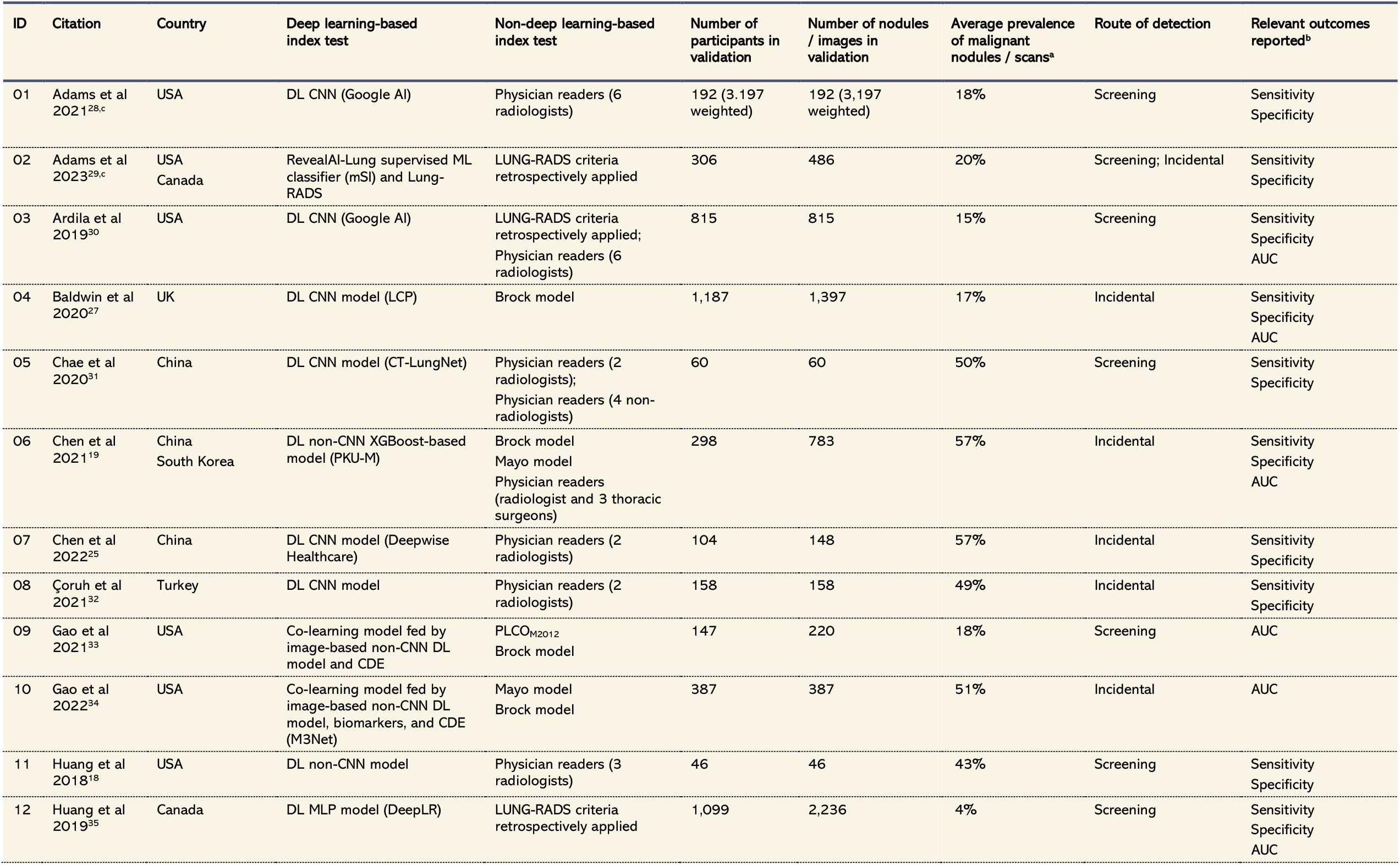
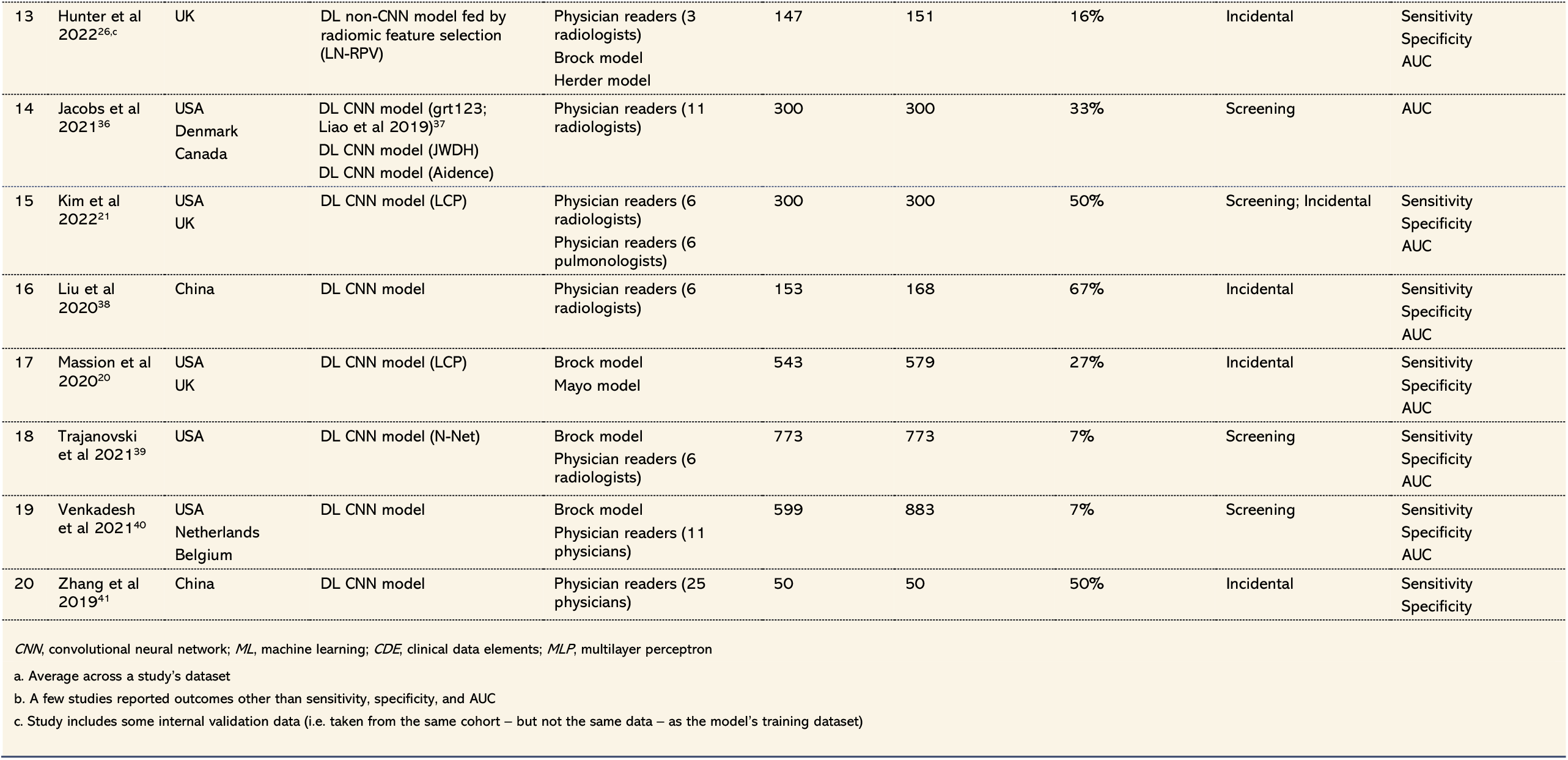
Characteristics of included studies.

| ID | Citation | Country | Deep learning-based index test | Non-deep learning-based index test | Number of participants in validation | Number of nodules / images in validation | Average prevalence of malignant nodules / scans<sup>a</sup> | Route of detection | Relevant outcomes reported<sup>b</sup> |
| --- | --- | --- | --- | --- | --- | --- | --- | --- | --- |
| 01 | Adams et al 2021<sup>28,c</sup> | USA | DL CNN (Google AI) | Physician readers (6 radiologists) | 192 (3,197 weighted) | 192 (3,197 weighted) | 18% | Screening | Sensitivity<br>Specificity |
| 02 | Adams et al 2023<sup>29,c</sup> | USA<br>Canada | RevealAI-Lung supervised ML classifier (mSI) and Lung-RADS | LUNG-RADS criteria retrospectively applied | 306 | 486 | 20% | Screening; Incidental | Sensitivity<br>Specificity |
| 03 | Ardila et al 2019<sup>30</sup> | USA | DL CNN (Google AI) | LUNG-RADS criteria retrospectively applied;<br>Physician readers (6 radiologists) | 815 | 815 | 15% | Screening | Sensitivity<br>Specificity<br>AUC |
| 04 | Baldwin et al 2020<sup>27</sup> | UK | DL CNN model (LCP) | Brock model | 1,187 | 1,397 | 17% | Incidental | Sensitivity<br>Specificity<br>AUC |
| 05 | Chae et al 2020<sup>31</sup> | China | DL CNN model (CT-LungNet) | Physician readers (2 radiologists);<br>Physician readers (4 non-radiologists) | 60 | 60 | 50% | Screening | Sensitivity<br>Specificity |
| 06 | Chen et al 2021<sup>19</sup> | China<br>South Korea | DL non-CNN XGBoost-based model (PKU-M) | Brock model<br>Mayo model<br>Physician readers (radiologist and 3 thoracic surgeons) | 298 | 783 | 57% | Incidental | Sensitivity<br>Specificity<br>AUC |
| 07 | Chen et al 2022<sup>25</sup> | China | DL CNN model (Deepwise Healthcare) | Physician readers (2 radiologists) | 104 | 148 | 57% | Incidental | Sensitivity<br>Specificity |
| 08 | Çoruh et al 2021<sup>32</sup> | Turkey | DL CNN model | Physician readers (2 radiologists) | 158 | 158 | 49% | Incidental | Sensitivity<br>Specificity |
| 09 | Gao et al 2021<sup>33</sup> | USA | Co-learning model fed by image-based non-CNN DL model and CDE | PLCO<sub>M2012</sub><br>Brock model | 147 | 220 | 18% | Screening | AUC |
| 10 | Gao et al 2022<sup>34</sup> | USA | Co-learning model fed by image-based non-CNN DL model, biomarkers, and CDE (M3Net) | Mayo model<br>Brock model | 387 | 387 | 51% | Incidental | AUC |
| 11 | Huang et al 2018<sup>18</sup> | USA | DL non-CNN model | Physician readers (3 radiologists) | 46 | 46 | 43% | Screening | Sensitivity<br>Specificity |
| 12 | Huang et al 2019<sup>35</sup> | Canada | DL MLP model (DeepLR) | LUNG-RADS criteria retrospectively applied | 1,099 | 2,236 | 4% | Screening | Sensitivity<br>Specificity<br>AUC |
| 13 | Hunter et al 2022<sup>26,c</sup> | UK | DL non-CNN model fed by radiomic feature selection (LN-RPV) | Physician readers (3 radiologists)<br>Brock model<br>Herder model | 147 | 151 | 16% | Incidental | Sensitivity<br>Specificity<br>AUC |
| 14 | Jacobs et al 2021<sup>36</sup> | USA<br>Denmark<br>Canada | DL CNN model (grt123; Liao et al 2019)<sup>37</sup><br>DL CNN model (JWDH)<br>DL CNN model (Aidence) | Physician readers (11 radiologists) | 300 | 300 | 33% | Screening | AUC |
| 15 | Kim et al 2022<sup>21</sup> | USA<br>UK | DL CNN model (LCP) | Physician readers (6 radiologists)<br>Physician readers (6 pulmonologists) | 300 | 300 | 50% | Screening; Incidental | Sensitivity<br>Specificity<br>AUC |
| 16 | Liu et al 2020<sup>38</sup> | China | DL CNN model | Physician readers (6 radiologists) | 153 | 168 | 67% | Incidental | Sensitivity<br>Specificity<br>AUC |
| 17 | Massion et al 2020<sup>20</sup> | USA<br>UK | DL CNN model (LCP) | Brock model<br>Mayo model | 543 | 579 | 27% | Incidental | Sensitivity<br>Specificity<br>AUC |
| 18 | Trajanovski et al 2021<sup>39</sup> | USA | DL CNN model (N-Net) | Brock model<br>Physician readers (6 radiologists) | 773 | 773 | 7% | Screening | Sensitivity<br>Specificity<br>AUC |
| 19 | Venkadesh et al 2021<sup>40</sup> | USA<br>Netherlands<br>Belgium | DL CNN model | Brock model<br>Physician readers (11 physicians) | 599 | 883 | 7% | Screening | Sensitivity<br>Specificity<br>AUC |
| 20 | Zhang et al 2019<sup>41</sup> | China | DL CNN model | Physician readers (25 physicians) | 50 | 50 | 50% | Incidental | Sensitivity<br>Specificity |
| *CNN*, convolutional neural network; *ML*, machine learning; *CDE*, clinical data elements; *MLP*, multilayer perceptron<br>a. Average across a study's dataset<br>b. A few studies reported outcomes other than sensitivity, specificity, and AUC<br>c. Study includes some internal validation data (i.e. taken from the same cohort – but not the same data – as the model's training dataset) |  |  |  |  |  |  |  |  |  |

All the studies save two were retrospective cohorts, with one study containign a prospective-cohort dataset, and one case-control study.^18,19^ The studies spanned continents and regions, with datasets taken from populations in North America (12 studies), Europe (7 studies), and East Asia (5 studies) (Table 1). Six of the studies used datasets taken from more than one country.

The outcome types in the studies included primarily assessed diagnostic performance. Some studies reported clinical utility measures, such as diagnostic re-classification.^20,21^ However, due to the inconsistency in the outcomes reported, it was not possible to conduct a meta-analysis on clinical utility outcomes. The main outcomes sought were sensitivity and specificity, for which 17 of the 20 studies reported outcomes, and AUC, for which 13 studies reported outcomes (Table 1).

Nineteen DL-based CADx models were identified from the included studies. The commonest type of learning algorithm used in the DL-based models was a Convolutional Neural Network (CNN). Twelve of the 19 models and 13 of the 20 included studies used a CNN algorithm as the basis for their DL malignancy prediction score.

For the external validation index tests, the commonest comparator was physician readers (15 of 33 datasets, from 14 studies). The majority of these readers were radiologists with ≥3 years’ experience. Among the non-radiologists, thoracic surgeons comprised the majority.

With the clinical risk models, the Brock model was the commonest validation method (12 datasets from eight studies), followed by the Mayo model with six datasets from three studies. This accords somewhat with the clinical risk models used in clinical practice. The Mayo model is considered the most externally validated model,^22^ but the Brock model has been shown to perform better than the Mayo model in screening populations.^23,24^

The majority of studies considered participants in the 50–75 age bracket, with very few examples of younger participants. All studies included both female and male participants. For the five studies that conducted external validation on datasets from the US National Lung Screening Trial (NLST), participants were all current or former heavy smokers.

Two studies excluded calcified nodules, and two studies excluded GGNs.^20,21,25^ The studies spanned the range of nodule sizes, with one study focussing only on malignancy risk prediction for large nodules >15 mm.^26^

In terms of prevalence of nodular malignancy, datasets ranged from as low as 2% up to 67%, with an average prevalence across all datsasets of 21.0%. A number of studies adjusted their dataset populations so that the number of malignant nodules matched the number of benign nodules. Despite this matching, most incidentally detected nodule populations had prevalence at or higher than 20%, whereas most screening populations had prevalence under 20%. This is reflected in real-world populations, where screening populations tend to have lower rates of nodular malignancy as compared to nodules that have been incidentally detected.^3^

### Diagnostic performance

For the DL-based CADx models, sensitivity ranged from 0.37 (95% CI: 0.25–0.50) for a 0.98 (95% CI: 0.95–0.99) specificity,^20^ to 1.00 (95% CI: 0.98–1.00) for a 0.28 (95% CI: 0.26–0.31) specificity (Figure 2A and Figure 3A).^27^ Pooled receiver operating characteristic (ROC) analysis of all DL-based CADx model results gave a pooled AUC of 0.90 (95% CI: 0.87–0.92), sensitivity of 0.88 (95% CI: 0.82–0.93) and specificity of 0.77 (95% CI: 0.69–0.84) (Figure 2A, Figure 3A, and Figure 4A). Pooled studies had an I^2^ index of 95.93% (95% CI: 95.04– 96.83) for sensitivity and 99.09% (95% CI: 98.97–99.22), corresponding to very high statistical heterogeneity. The Deeks funnel plot showed no significant asymmetry, indicating no evidence of publication bias (eFigure 2).

**Figure 2.**
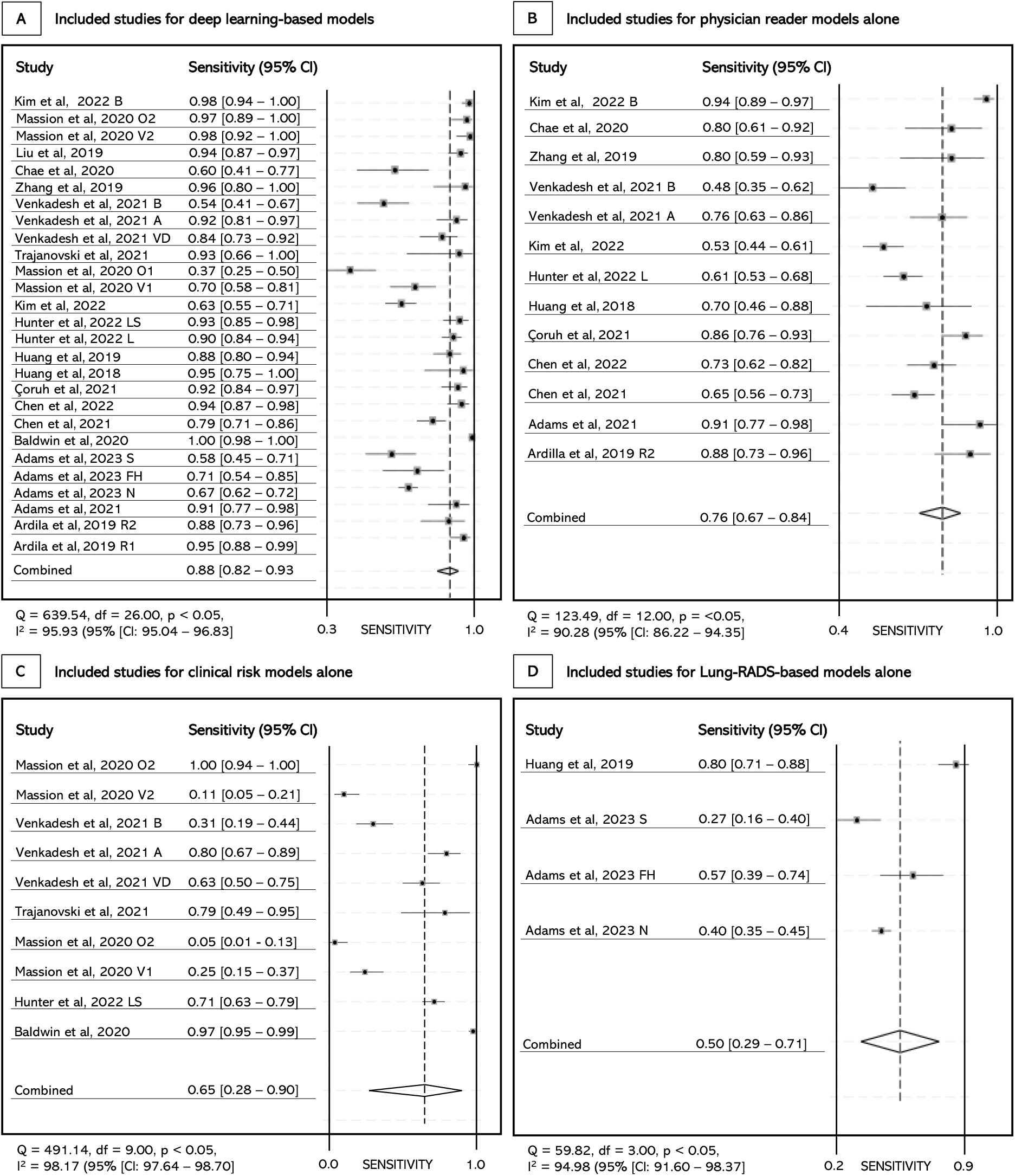
Pooled sensitivity analyses of the included studies and their datasets

**Figure 3.**
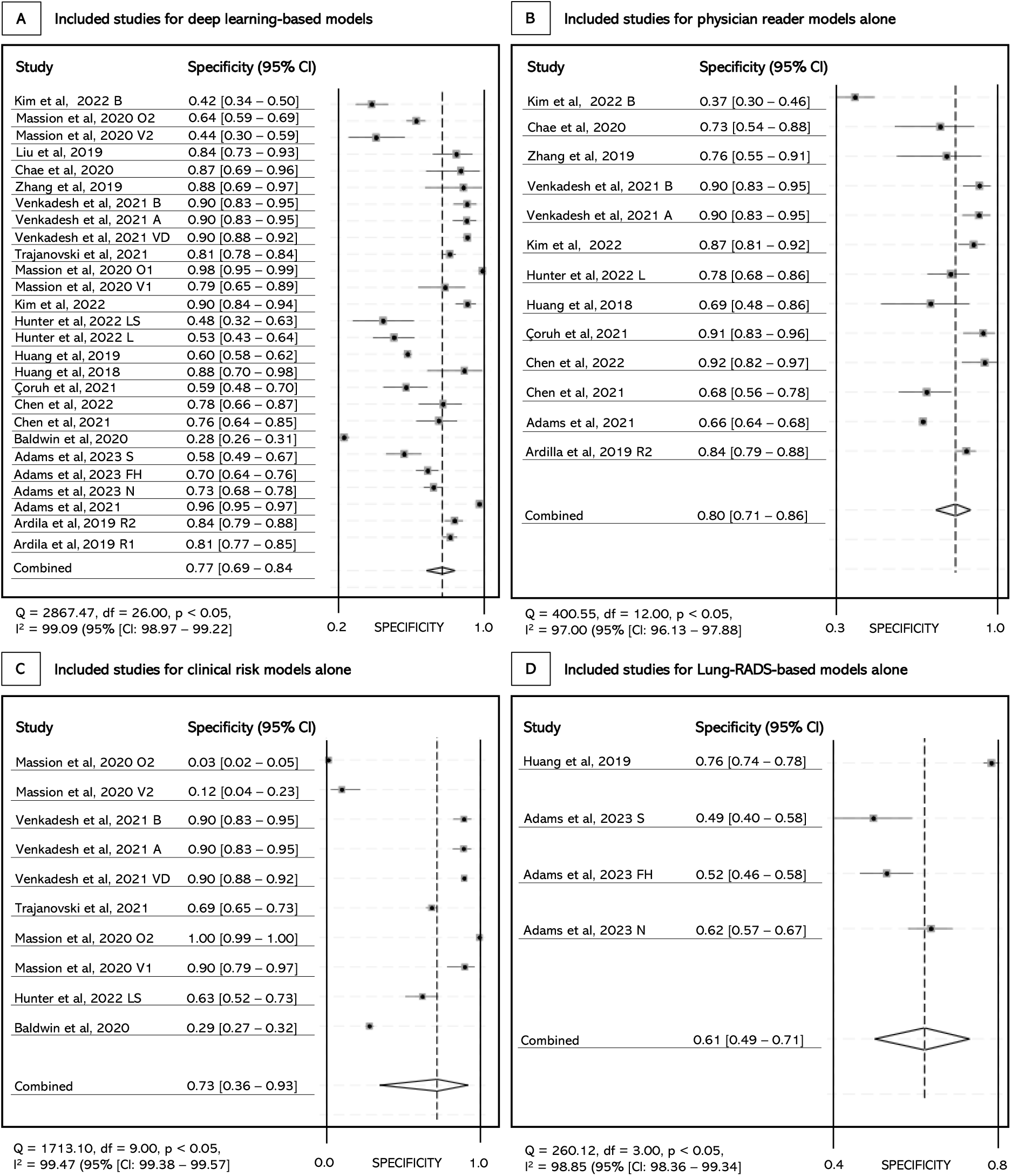
Pooled specificity analyses of the included studies and their datasets

**Figure 4.**
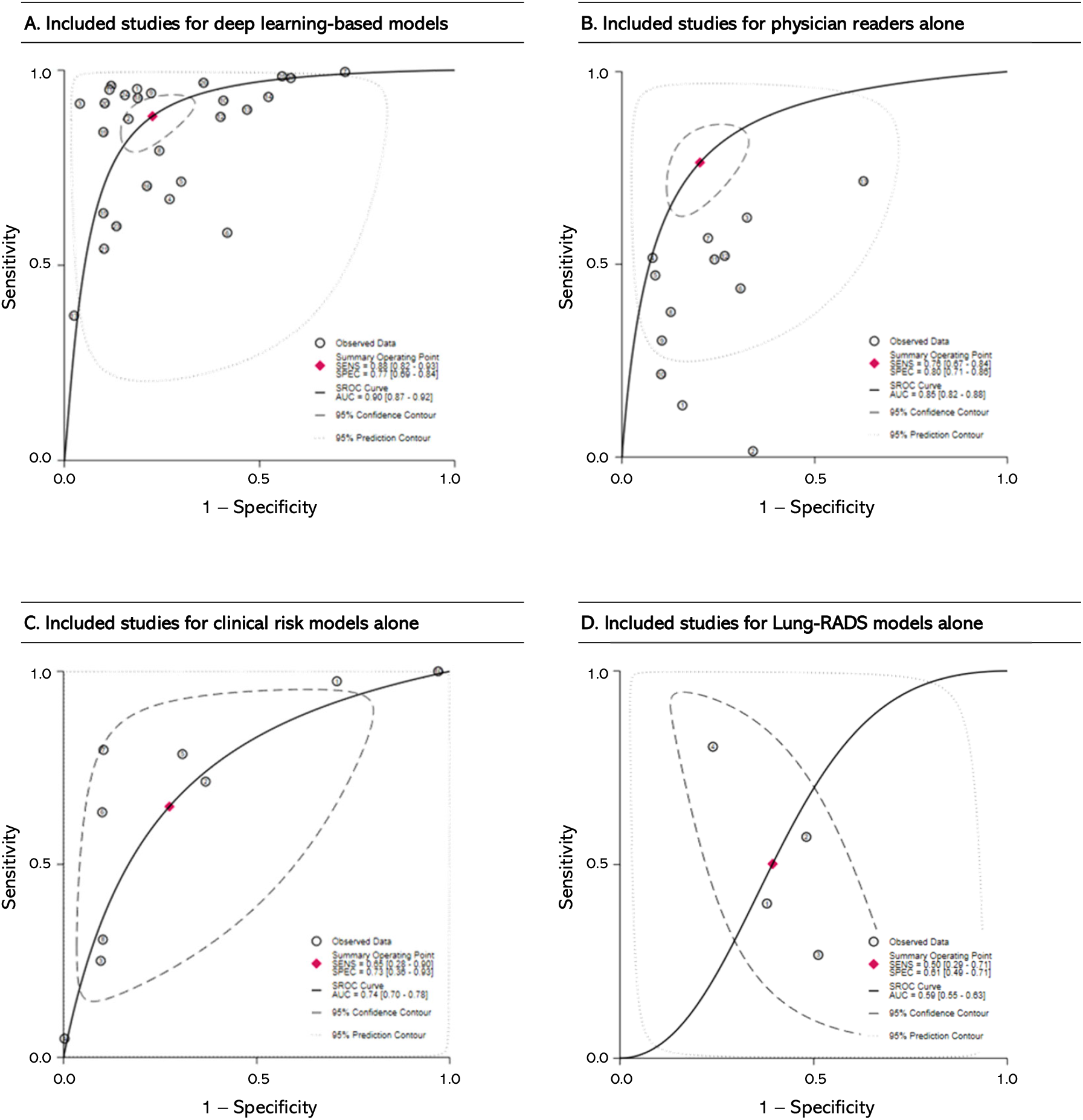
Summary ROC curve analyses of included diagnostic models

Separate pooled analysis for physician readers gave a pooled AUC of 0.85 (95% CI: 0.82–0.88), sensitivity of 0.76 (95% CI: 0.67–0.84) and specificity of 0.80 (95% CI: 0.71–0.86) (Figure 2B, Figure 3B, and Figure 4B). Pooled studies had an I^2^ index of 90.28% (95% CI: 86.22–94.35) for sensitivity and 97.00% (95% CI: 96.13– 97.88), indicating high statistical heterogeneity.

Pooled analysis for clinical risk models gave a pooled AUC of 0.74 (95% CI: 0.70–0.78), sensitivity of 0.65 (95% CI: 0.28–0.90) and specificity of 0.73 (95% CI: 0.36–0.93) (Figure 2C, Figure 3C, and Figure 4C). Pooled studies had an I^2^ index of 98.17% (95% CI: 97.64–98.70) for sensitivity and 99.47% (95% CI: 99.38–99.57), indicating very high statistical heterogeneity.

Lastly, pooled analysis for Lung-RADS-based models gave a pooled AUC of 0.59 (95% CI: 0.55–0.63), sensitivity of 0.50 (95% CI: 0.29–0.71) and specificity of 0.61 (95% CI: 0.49–0.71) (Figure 2D, Figure 3D, and Figure 4D).

Pooled studies had an I^2^ index of 94.98% (95% CI: 91.60–98.37) for sensitivity and 98.85% (95% CI: 98.36– 99.34), indicating very high statistical heterogeneity.

Sub-group analyses revealed that DL-based CADx models displayed significantly higher sensitivity on incidentally detected nodules than on screening-detected nodules, 0.91 (95% CI: 0.81–0.96) *versus* 0.84 (95% CI: 0.76– 0.90), respectively (eFigure 3). This increased reliability in detecting lung cancer came at the cost of specificity with screening-detected nodules having 0.84 (95% CI: 0.78–0.89) as compared to incidentally detected nodules at 0.70 (95% CI: 0.55–0.81). There was no significant difference for physician readers between screening and incidental detection. However, clinical risk models showed superior specificity for screening-detected nodules as compared to incidentally detected nodules, 0.86 (95% CI: 0.77–0.92) *versus* 0.59 (95% CI: 0.11–0.95), respectively (eFigure 4).

Further sub-group analyses were carried out on prevalence. To do this, we took the baseline prevalence of malignancy in CT-detected pulmonary nodules in the US, ∼5%,^3^ and multiplied it by a factor of 4, and used this as the threshold for classifying a study’s prevalence as high or low. Our reasoning was that if a study’s prevalence was 4 times as high as the baseline population prevalence, it could safely be considered high. Thus, our threshold for defining studies as having high or low prevalence was 20%. DL-based CADx models performed significantly better in low-prevalence studies than in high-prevalence studies: sensitivity of 0.90 (95% CI: 0.78–0.96) and specificity of 0.81 (95% CI: 0.66–0.90) as compared to sensitivity of 0.87 (95% CI: 0.78–0.92) and specificity of 0.75 (95% CI: 0.66–0.82), respectively.

### Quality assessment

The results of the quality assessment using QUADAS-2 are shown in eTable 2 in the Supplementary Material. Overall, a low-to-moderate risk of bias was found in most studies.

## Discussion

A systematic review and meta-analysis that investigated the diagnostic performance of DL-based CADx models in predicting the risk of malignancy in pulmonary nodules *versus* methods currently used in clinical practice was performed. DL-based CADx models were significantly more sensitive than physician judgement alone, 15.8% (0.88 *v* 0.76), and clinical risk models alone, 35.4% (0.88 *v* 0.65). They had approximately the same pooled specificity as physician judgement alone (0.77 [95% CI: 0.69 –0.84] *v* 0.80 [95% CI: 0.71 –0.86], respectively), but were 5.5% (0.77 *v* 0.73) more specific than clinical risk models alone.

Accounting for threshold effects, DL-based CADx models had significantly superior sAUROC, with relative sAUROCs of 1.06 (95% CI: 1.03–1.08), 1.22 (95% CI: 1.19–1.24), and 1.53 (95% CI: 1.50–1.55) as compared to physician judgement alone, clinical risk models alone, and Lung-RADS-based models alone, respectively.

This study attempted to exhaustively search the literature for all studies and models relevant to the research question. During screening, two of the commonest reasons for ineligibility were that the study did not conduct any direct external validation of the DL-based CADx model being analysed (28 studies with no direct external validation at the final screening stage) and that the study’s model was for detection of pulmonary nodules, not classification or diagnosis of them (11 studies with ineligible index tests at the final screening stage) (Figure 1).

In order to evaluate their performance when applied to different populations, it is crucial to conduct some external validation of DL-based risk prediction models in populations independent of those used during model development.^42^ The majority of studies identified were on development and wholly internal validation of the DL-based model or validation against other DL-based models, and not on external validation against models currently used in clinical practice.

On the second commonest exclusion, computer-aided solutions for pulmonary nodule management can be broadly categorised into two types: computer-aided detection (CADe) and CADx (diagnosis). CADe involves a module designed to detect suspicious lung nodules and segment them for further analysis. Its purpose is to assist in the identification of potential abnormalities. CADx, on the other hand, goes beyond detection. It provides a nodule-level and, possibly, patient-level classification of the risk of malignancy. Only CADx is considered in this systematic review and meta-analysis. In broad terms, detection of pulmonary nodules is relatively easy. Distinguishing malign nodules from benign ones is not.^2,5,43^

Two previous systematic reviews have studied this issue,^11,12^ albeit without the direct comparison between DL-based CADx models and external validation with methods used in clinical practice. Forte et al 2022 was the only one to conduct a meta-analysis, considering six studies, all of which are also included in this review and analysis. Pooled sensitivity and specificity were 0.94 (95% CI: 0.86–0.98) and 0.69 (95% CI: 0.51–0.83), respectively, both with significant heterogeneity, while sAUROC was 0.90 (95% CI: 0.86–0.92).^11^ No quantitative comparisons against physician reader or clinical risk models alone were performed, and nor were any sub-group analyses performed. The authors noted that DL-based CADx models performed well and that as non-invasive methods, they could provide support to clinics in detecting and diagnosing lung cancer early.

### Limitations

Although these results strongly support the use of externally validated DL-based CADx models, two primary limitations were noted. Only observational studies could be found, and of these only one was prospective. No randomised controlled trials or other interventional studies were found. This is to be expected given that evidentiary requirements for diagnostic tools are not set as high as therapeutic interventions (drugs and biologics), and the difficulty in conducting such studies with diagnostic tools.^44^ The second limitation was the high heterogeneity found among studies.

Sources of heterogeneity were investigated by conducting sub-group analyses. Prevalence was found to be a weak source of heterogeneity for clinical risk models in particular. Age range of the study population was another moderate source of heterogeneity across all models. However, the strongest source of heterogeneity was likely the threshold or operating cut-off point used by researchers in testing the models. The types of thresholds used varied considerably from study to study. They included fixing the specificity of models to 0.90,^40^ to setting rule-out (definite benignity) malignancy scores at 0.05 (out of 1.0) or rule-in (definite malignancy) malignancy scores to 0.65 (out of 1.0).^20,21^ Sensitivity to threshold effects was not investigated due to these inconsistent methods. However, the inclusion of AUC and its concordance with the sensitivity and specificity for each index test type helped alleviate this concern. Additionally, the low-to-moderate risk of bias found in most studies, and no significant publication bias demonstrated the findings were robust, in spite of the high heterogeneity.

Placing it further in context, the high heterogeneity in DL-based CADx models makes sense given the very different models under consideration, and the further work required on calibrating these models. However, as more validation in clinical practice occurs, and certain models become standard use in clinical practice – as has happened with the Mayo and Brock models for clinical risk models – heterogeneity may reduce.^7,8^ On this point, it important to note that use in clinical practice is important. Other clinical risk models, such as the Gurney model and the Bayesian Inference Malignancy Calculator (BIMC),^45–47^ both of which use Bayesian analysis of clinico-demographic variables rather than logistic regression, have undergone external validation but are not used in clinical practice. Taking excellent diagnostic performance into clinical practice for further calibration and validation is the next step to ensuring improved patient outcomes are fully realised.

## Conclusion

These results demonstrate that DL-based CADx models have superior or comparable diagnostic performance as compared to methods currently used in clinical practice. The results support the use of DL-based CADx models alongside physician readers in clinical practice, especially for the management of incidentally detected nodules. While further research is required before they become an essential and routine part of the physician’s toolkit, recommendation for use in clinical practice as an option in the physician’s tookit is justified by our findings.

## Data Availability

All data produced in the present study are available upon reasonable request to the authors

## References

1. Woodard GA, Jones KD, Jablons DM. Lung Cancer Staging and Prognosis. Cancer Treat Res. 2016;170:47–75. doi:10.1007/978-3-319-40389-2_3

2. Loverdos K, Fotiadis A, Kontogianni C, Iliopoulou M, Gaga M. Lung nodules: A comprehensive review on current approach and management. Ann Thorac Med. 2019;14(4):226–238. doi:10.4103/atm.ATM_110_19

3. Mazzone PJ, Lam L. Evaluating the Patient With a Pulmonary Nodule. JAMA. 2022;327(3):264. doi:10.1001/jama.2021.24287

4. Mahesh M, Ansari AJ, Mettler FA. Patient Exposure from Radiologic and Nuclear Medicine Procedures in the United States and Worldwide: 2009–2018. Radiology. 2023;307(1). doi:10.1148/radiol.221263

5. Paez R, Kammer MN, Massion P. Risk stratification of indeterminate pulmonary nodules. Curr Opin Pulm Med. 2021;27(4):240–248. doi:10.1097/MCP.0000000000000780

6. Peikert T, Bartholmai BJ, Maldonado F. Radiomics-based Management of Indeterminate Lung Nodules? Are We There Yet? Am J Respir Crit Care Med. 2020;202(2):165–167. doi:10.1164/rccm.202004-1279ED

7. Swensen SJ, Silverstein MD, Ilstrup DM, Schleck CD, Edell ES. The probability of malignancy in solitary pulmonary nodules. Application to small radiologically indeterminate nodules. Arch Intern Med. 1997;157(8):849–855.

8. McWilliams A, Tammemagi MC, Mayo JR, et al. Probability of Cancer in Pulmonary Nodules Detected on First Screening CT. New England Journal of Medicine. 2013;369(10):910–919. doi:10.1056/NEJMoa1214726

9. Li Y, Wang J. A mathematical model for predicting malignancy of solitary pulmonary nodules. World J Surg. 2012;36(4):830–835. doi:10.1007/s00268-012-1449-8

10. Lee JH, Hwang EJ, Kim H, Park CM. A narrative review of deep learning applications in lung cancer research: from screening to prognostication. Transl Lung Cancer Res. 2022;11(6):1217–1229. doi:10.21037/tlcr-21-1012

11. Forte GC, Altmayer S, Silva RF, et al. Deep Learning Algorithms for Diagnosis of Lung Cancer: A Systematic Review and Meta-Analysis. Cancers (Basel). 2022;14(16). doi:10.3390/cancers14163856

12. Wu Z, Wang F, Cao W, et al. Lung cancer risk prediction models based on pulmonary nodules: A systematic review. Thorac Cancer. 2022;13(5):664–677. doi:10.1111/1759-7714.14333

13. Page MJ, McKenzie JE, Bossuyt PM, et al. The PRISMA 2020 statement: an updated guideline for reporting systematic reviews. BMJ. Published online March 29, 2021:n71. doi:10.1136/bmj.n71

14. Chelala L, Hossain R, Kazerooni EA, Christensen JD, Dyer DS, White CS. Lung-RADS Version 1.1: Challenges and a Look Ahead, From the AJR Special Series on Radiology Reporting and Data Systems. American Journal of Roentgenology. 2021;216(6):1411–1422. doi:10.2214/AJR.20.24807

15. Whiting PF, Rutjes AWS, Westwood ME, et al. QUADAS-2: a revised tool for the quality assessment of diagnostic accuracy studies. Ann Intern Med. 2011;155(8):529–536. doi:10.7326/0003-4819-155-8-201110180-00009

16. StataCorp LLC. Stata Statistical Software: Release 18. Published online 2023.

17. The Cochrane Collaboration. Review Manager (RevMan). Published online 2020. Accessed June 6, 2023. http://www.revman.cochrane.org

18. Huang P, Park S, Yan R, et al. Added Value of Computer-aided CT Image Features for Early Lung Cancer Diagnosis with Small Pulmonary Nodules: A Matched Case-Control Study. Radiology. 2018;286(1):286–295. doi:10.1148/radiol.2017162725

19. Chen K, Nie Y, Park S, et al. Development and Validation of Machine Learning-based Model for the Prediction of Malignancy in Multiple Pulmonary Nodules: Analysis from Multicentric Cohorts. Clin Cancer Res. 2021;27(8):2255–2265. doi:10.1158/1078-0432.CCR-20-4007

20. Massion PP, Antic S, Ather S, et al. Assessing the Accuracy of a Deep Learning Method to Risk Stratify Indeterminate Pulmonary Nodules. Am J Respir Crit Care Med. 2020;202(2):241–249. doi:10.1164/rccm.201903-0505OC

21. Kim RY, Oke JL, Pickup LC, et al. Artificial Intelligence Tool for Assessment of Indeterminate Pulmonary Nodules Detected with CT. Radiology. 2022;304(3):683–691. doi:10.1148/radiol.212182

22. Choi HK, Ghobrial M, Mazzone PJ. Models to Estimate the Probability of Malignancy in Patients with Pulmonary Nodules. Ann Am Thorac Soc. 2018;15(10):1117–1126. doi:10.1513/AnnalsATS.201803-173CME

23. González Maldonado S, Delorme S, Hüsing A, et al. Evaluation of Prediction Models for Identifying Malignancy in Pulmonary Nodules Detected via Low-Dose Computed Tomography. JAMA Netw Open. 2020;3(2):e1921221. doi:10.1001/jamanetworkopen.2019.21221

24. White CS, Dharaiya E, Campbell E, Boroczky L. The Vancouver Lung Cancer Risk Prediction Model: Assessment by Using a Subset of the National Lung Screening Trial Cohort. Radiology. 2017;283(1):264–272. doi:10.1148/radiol.2016152627

25. Chen Y, Tian X, Fan K, Zheng Y, Tian N, Fan K. The Value of Artificial Intelligence Film Reading System Based on Deep Learning in the Diagnosis of Non-Small-Cell Lung Cancer and the Significance of Efficacy Monitoring: A Retrospective, Clinical, Nonrandomized, Controlled Study. Comput Math Methods Med. 2022;2022:2864170. doi:10.1155/2022/2864170

26. Hunter B, Chen M, Ratnakumar P, et al. A radiomics-based decision support tool improves lung cancer diagnosis in combination with the Herder score in large lung nodules. EBioMedicine. 2022;86:104344. doi:10.1016/j.ebiom.2022.104344

27. Baldwin DR, Gustafson J, Pickup L, et al. External validation of a convolutional neural network artificial intelligence tool to predict malignancy in pulmonary nodules. Thorax. 2020;75(4):306–312. doi:10.1136/thoraxjnl-2019-214104

28. Adams SJ, Mondal P, Penz E, Tyan CC, Lim H, Babyn P. Development and Cost Analysis of a Lung Nodule Management Strategy Combining Artificial Intelligence and Lung-RADS for Baseline Lung Cancer Screening. Journal of the American College of Radiology. 2021;18(5):741–751. doi:10.1016/j.jacr.2020.11.014

29. Adams SJ, Madtes DK, Burbridge B, et al. Clinical Impact and Generalizability of a Computer-Assisted Diagnostic Tool to Risk-Stratify Lung Nodules With CT. Journal of the American College of Radiology. 2023;20(2):232–242. doi:10.1016/j.jacr.2022.08.006

30. Ardila D, Kiraly AP, Bharadwaj S, et al. End-to-end lung cancer screening with three-dimensional deep learning on low-dose chest computed tomography. Nat Med. 2019;25(6):954–961. doi:10.1038/s41591-019-0447-x

31. Chae KJ, Jin GY, Ko SB, et al. Deep Learning for the Classification of Small (≤2 cm) Pulmonary Nodules on CT Imaging: A Preliminary Study. Acad Radiol. 2020;27(4):e55–e63. doi:10.1016/j.acra.2019.05.018

32. Gürsoy Çoruh A, Yenigün B, Uzun Ç, et al. A comparison of the fusion model of deep learning neural networks with human observation for lung nodule detection and classification. Br J Radiol. 2021;94(1123):20210222. doi:10.1259/bjr.20210222

33. Gao R, Tang Y, Khan MS, et al. Cancer Risk Estimation Combining Lung Screening CT with Clinical Data Elements. Radiol Artif Intell. 2021;3(6):e210032. doi:10.1148/ryai.2021210032

34. Gao R, Li T, Tang Y, et al. Reducing uncertainty in cancer risk estimation for patients with indeterminate pulmonary nodules using an integrated deep learning model. Comput Biol Med. 2022;150:106113. doi:10.1016/j.compbiomed.2022.106113

35. Huang P, Lin CT, Li Y, et al. Prediction of lung cancer risk at follow-up screening with low-dose CT: a training and validation study of a deep learning method. Lancet Digit Health. 2019;1(7):e353–e362. doi:10.1016/S2589-7500(19)30159-1

36. Jacobs C, Setio AAA, Scholten ET, et al. Deep Learning for Lung Cancer Detection on Screening CT Scans: Results of a Large-Scale Public Competition and an Observer Study with 11 Radiologists. Radiol Artif Intell. 2021;3(6):e210027. doi:10.1148/ryai.2021210027

37. Liao F, Liang M, Li Z, Hu X, Song S. Evaluate the Malignancy of Pulmonary Nodules Using the 3-D Deep Leaky Noisy-OR Network. IEEE Trans Neural Netw Learn Syst. 2019;30(11):3484–3495. doi:10.1109/TNNLS.2019.2892409

38. Liu J, Zhao L, Han X, Ji H, Liu L, He W. Estimation of malignancy of pulmonary nodules at CT scans: Effect of computer-aided diagnosis on diagnostic performance of radiologists. Asia Pac J Clin Oncol. 2021;17(3):216–221. doi:10.1111/ajco.13362

39. Trajanovski S, Mavroeidis D, Swisher CL, et al. Towards radiologist-level cancer risk assessment in CT lung screening using deep learning. Comput Med Imaging Graph. 2021;90:101883. doi:10.1016/j.compmedimag.2021.101883

40. Venkadesh KV, Setio AAA, Schreuder A, et al. Deep Learning for Malignancy Risk Estimation of Pulmonary Nodules Detected at Low-Dose Screening CT. Radiology. 2021;300(2):438–447. doi:10.1148/radiol.2021204433

41. Zhang C, Sun X, Dang K, et al. Toward an Expert Level of Lung Cancer Detection and Classification Using a Deep Convolutional Neural Network. Oncologist. 2019;24(9):1159–1165. doi:10.1634/theoncologist.2018-0908

42. Ramspek CL, Jager KJ, Dekker FW, Zoccali C, van Diepen M. External validation of prognostic models: what, why, how, when and where? Clin Kidney J. 2021;14(1):49–58. doi:10.1093/ckj/sfaa188

43. Maldonado F, Lentz RJ. Reducing uncertainty to a manageable level: the need for a nuanced and patient-centric approach to lung nodule management in the 21st century. J Thorac Dis. 2020;12(6):3242–3244. doi:10.21037/jtd.2020.03.65

44. Mazumdar M, Zhong X, Ferket B. Diagnostic Trials. In: Principles and Practice of Clinical Trials. Springer International Publishing; 2021:1–28. doi:10.1007/978-3-319-52677-5_281-1

45. Gurney JW. Determining the likelihood of malignancy in solitary pulmonary nodules with Bayesian analysis. Part I. Theory. Radiology. 1993;186(2):405–413. doi:10.1148/radiology.186.2.8421743

46. Soardi GA, Perandini S, Motton M, Montemezzi S. Assessing probability of malignancy in solid solitary pulmonary nodules with a new Bayesian calculator: improving diagnostic accuracy by means of expanded and updated features. Eur Radiol. 2015;25(1):155–162. doi:10.1007/s00330-014-3396-2

47. Gurney JW, Lyddon DM, McKay JA. Determining the likelihood of malignancy in solitary pulmonary nodules with Bayesian analysis. Part II. Application. Radiology. 1993;186(2):415–422. doi:10.1148/radiology.186.2.8421744

